# How often did syphilis tests have corresponding HIV tests in Ontario, Canada? A retrospective analysis of comprehensive laboratory data

**DOI:** 10.1101/2025.07.06.25330981

**Authors:** Sean Colyer, Ellen G. Avery, Abigail Kroch, Juan Liu, Austin Zygmunt, Maya Kesler, Ashleigh Sullivan, Vanessa Tran

## Abstract

**Objectives:** Canadian guidelines recommend HIV testing for individuals being evaluated for syphilis. Our objective was to examine three aspects of HIV testing (i.e., if a HIV test occurred, timing of HIV test in relation to syphilis test, and the proportion with a positive HIV test result) among syphilis tests between 2017 and 2022 from individuals with no evidence of a previous HIV diagnosis.

**Design and setting:** This study is a retrospective analysis of comprehensive laboratory testing data from Ontario’s provincial public health laboratory.

**Participants:** Direct fluorescent antibody (DFA) and serological non-prenatal syphilis tests from January 1, 2017 to December 31, 2022, from individuals aged ≥15 years with no evidence of a previous HIV diagnosis (N = 3,001,058 total tests). Positive syphilis tests were categorized using the rapid plasma reagin (RPR) titre as “current” (DFA+/RPR ≥1:8) or “historical” (RPR <1:8). Exposure categories were assigned using individually-linked HIV exposure category data retrieved from the laboratory’s HIV database.

**Primary and secondary outcome measures:** Number and proportion of syphilis tests that had a corresponding HIV test on the same day or within 7/28/90/180 days, and among those with an HIV test within 28 days, number and proportion that had an HIV-positive test result.

**Results:** From 2017 - 2022, 1,516,726 and 1,484,332 syphilis tests among males and females, respectively, were included in the analysis. Individuals with a positive syphilis result were less likely to be tested for HIV within 28 days of their syphilis test compared to those with a negative syphilis test result (74.7% versus 91.1% in males, 97.5% CI [-0.17, -0.16], 65.2% versus 92.4% in females, 97.5% CI [-0.28, -0.26]). Males with “current” positive syphilis test results were less likely than males with “historical” positive syphilis results to be tested for HIV within 28 days (69.1% versus 76.6%, 97.5% CI [-0.084, -0.066]); this was not true in females (67.1% versus 64.4%, 97.5% CI [0.0062, 0.049]). Males overall and males with “current” syphilis were more likely to be diagnosed as HIV-positive (p<0.025).

**Conclusions:** Most individuals who tested for syphilis at PHO were also tested for HIV, though those positive for syphilis were less likely to be tested, representing an opportunity for enhanced HIV testing. Ensuring that syphilis-positive individuals are tested for HIV may identify previously undiagnosed individuals living with HIV.

## MAIN TEXT

### STRENGTHS AND LIMITATIONS

- As the provincial public health laboratory conducts nearly all syphilis and HIV diagnostic and HIV viral load testing in the province, our data represents a comprehensive analysis of our research questions, and our results are representative of the province.
- Individuals’ HIV tests may not always link to their syphilis tests. This has implications for [1] identifying people previously diagnosed with HIV (and exclusion of their syphilis tests); and [2] identifying corresponding HIV tests. Although we employed linkage based on health card numbers (a unique ID for most Ontarians), dates of birth, and first and last names, failed linkages could result from incomplete or inconsistent data, or non-nominal HIV testing. The majority of anonymous HIV testing in Ontario is conducted in males (20), therefore males may be more likely to have no linked HIV test.
- Given that we were limited to syphilis laboratory testing data alone, we could not distinguish current syphilis infections from treated/resolved infections. Our “current” definition (i.e. RPR ≥1:8) is a conservative estimate of infectious syphilis that was based on national guidelines and consultation with health care providers with syphilis expertise (5). Our categorization based on RPR titres is not a validated method, and will encompass misclassification on the individual level. Further, the lack of longitudinal analyses also contributes to the misclassification at the individual level. However, we believe that our simplified method applied at the population level is sufficient to address the questions.
- We had incomplete data with respect to HIV exposure category, and these data are subject to recall and social desirability biases, as discussed. This may affect interpretations among exposure categories.

## INTRODUCTION

Despite decreases in HIV testing and diagnosis rates in 2020 and 2021 in Ontario, Canada related to the COVID-19 pandemic, these returned to near pre-pandemic levels in 2022 (1). Consequently, along with other interventions, timely and prudent testing for HIV remains an important secondary prevention measure. Concurrent with the robust rates of HIV diagnoses have been increasing rates of infectious syphilis, which, with the exception of 2020, have increased steadily and dramatically (>3-fold) in Ontario between 2013 and 2022 (2). This rise in syphilis cases, and increased syphilis screening and diagnostic testing associated with this increase, represent opportunities for identification and testing of individuals at elevated HIV risk.

Guidelines from the Public Health Agency of Canada (PHAC) recommend screening for syphilis for those with new or multiple sex partners or upon request (3). The association between a bacterial sexually transmitted infection (STI) diagnosis and subsequent HIV acquisition is well documented (4). As such, PHAC guidelines recommend that people being evaluated or treated for syphilis be screened for HIV (5), while Ontario’s HIV testing guidelines recommend HIV testing for members of populations with higher HIV rates when an STI is diagnosed (6). PHAC guidelines further recommend that those at risk for HIV be screened at least annually (7). The United States Center for Disease Control and Prevention (CDC) similarly state that HIV testing should be performed concurrently with testing for other STIs (8). Because of the high correlation between STIs, especially syphilis, and HIV acquisition, the CDC guidelines also state that if the initial HIV testing opportunity is missed, a STI diagnosis should be used as an opportunity to test for HIV (8). Given the epidemiology of HIV and syphilis and their respective testing guidelines, individuals testing for syphilis represent an informative population to investigate HIV testing efforts. HIV testing frequencies can also be compared to published guidelines to facilitate assessment of recommended HIV testing.

Variability exists in HIV risk following an STI diagnosis, with gay, bisexual, and other men who have sex with men (GBMSM) at elevated risk. One study in New York City demonstrated that 1 in 20 GBMSM were diagnosed with HIV within a year of their syphilis diagnosis (9). In a Danish GBMSM cohort, a new syphilis diagnosis was associated with a 9.8% risk of acquiring HIV over the subsequent five years (10). In a cohort of men in Baltimore, while an STI diagnosis increased the risk for a subsequent HIV diagnosis in all men, GBMSM had a much higher HIV incidence than non-GBMSM (11). Less is known about the HIV acquisition risk in those that receive negative STI test results. Though HIV incidence and prevalence are broadly lower in women than men in Canada (12) (13), the frequency of HIV screening in women who have been tested for syphilis is unknown. Surveillance data in Manitoba, Canada recently demonstrated increased social vulnerability in newly HIV-diagnosed women compared to newly HIV-diagnosed men, suggesting that at-risk women may be harder for prevention efforts to reach (14).

In Ontario, testing for syphilis and HIV is performed almost exclusively at Public Health Ontario (PHO)’s laboratory, which is the provincial public health reference laboratory. It is unclear whether individuals in the province are being screened for both infectious diseases as per the aforementioned guidelines, or whether gaps exist in screening practices, as has been demonstrated elsewhere (15). Using laboratory testing data, our objective was to examine three aspects of HIV testing among syphilis tests performed between 2017 and 2022 from individuals with no evidence of a previous HIV diagnosis: [1] to determine if an HIV test occurred; [2] the timing of the HIV test in relation to syphilis test; and [3] the proportion with a positive HIV test result. Our comprehensive dataset allows for the assessment of HIV testing practices over different time windows (e.g. within 28 to 180 days following a syphilis diagnosis), which enables us to determine whether STI screening is occurring in Ontario as recommended. To the best of our knowledge, this is the first time this type of analysis has been done at the provincial level.

## METHODS

The vast majority of syphilis screening and all syphilis confirmatory testing in Ontario is centralized at Public Health Ontario. Syphilis serology and direct fluorescent antibody (DFA) testing data were extracted from PHO’s laboratory information management system (LIMS), for tests from January 1, 2017 to December 31, 2022. The following syphilis tests were excluded: quality control tests; prenatal tests; tests from individuals aged <15 years (and unknown date of birth); duplicate tests from the same individual on the same date.

HIV testing in Ontario is centralized at Public Health Ontario except for rare exceptions. As a result, the HIV DataMart contains nearly all HIV diagnostic and viral load testing conducted in Ontario. HIV testing data (HIV serology and point-of-care test result, HIV exposure category, identifying information) were extracted from PHO’s 2022 HIV DataMart, from the date of when data were first collected (1985 onwards for HIV diagnostic testing, 1996 onwards for HIV viral load testing) to December 31, 2022. Syphilis tests and HIV tests from the same individual were linked via matching health card numbers, or if missing, first name, last name, and date of birth (exact match). Since the purpose of HIV screening is to find new HIV diagnoses, we restricted syphilis tests to individuals with no evidence of a previous HIV diagnosis as of the date of the syphilis test. To do this we excluded: [1] all syphilis tests from an individual who reported being previously diagnosed with HIV (via the HIV test requisition or a follow-up questionnaire sent to the ordering physician of HIV-positive tests); [2] all syphilis tests occurring after an individual’s first positive diagnostic HIV test; and [3] all syphilis tests occurring on the same date or after an individual’s first HIV viral load test. For each syphilis test, the number of days between the date of the syphilis test and the most proximal-in-time HIV test (either preceding or following) was calculated. This was used to categorize syphilis tests with respect to timing of “corresponding” HIV test (i.e. same day (i.e. zero days), same day +/-28 days, etc). Similarly, the number of days to the patient’s subsequent positive HIV test (if any) was calculated for each syphilis test (note that the number of days following a previous positive HIV test was not calculated, as these syphilis tests would have been excluded for already being HIV-diagnosed per above). The date the specimen was received/logged in the laboratory was used as test date instead of collection date because the latter is inconsistently provided. The analysis was completed on the test level, as opposed to the individual level, with individuals having multiple syphilis tests included in the analysis.

Given that we were limited to syphilis laboratory testing data alone (i.e. no clinical information), and that an individual is likely to always screen positive for syphilis once infected, we aimed to provide a population-level approximation of current or active versus historical syphilis infections. We created two categories of positive syphilis tests: [1] “Current”: DFA-positive or a positive chemiluminescent microparticle immunoassay (CMIA) serology screen with a rapid plasma regain (RPR) antibody titre of

≥1:8 were considered more likely to represent current syphilis infections; and [2] “Historical”: positive CMIA screen with an RPR <1:8 were considered more likely to represent historical (treated/resolved) infections. All other syphilis tests were considered “non-positive” tests (including negative, inconclusive, and invalid results). Invalid results were retained as they still represent instances of syphilis tests being ordered/requested.

HIV laboratory surveillance data includes collection of HIV risk factors on the test requisition form and a follow-up questionnaire post diagnosis to create HIV exposure categories; Ontario’s syphilis testing data does not include this information (16). To assign exposure categories to syphilis tests, individually-linked HIV exposure data were used. As this information is not reported for the majority (>70%) of HIV tests (17), HIV risk factors reported on earlier or later HIV tests (whichever was closest in time) were used to assign exposure categories for the syphilis tests that had corresponding HIV tests. In doing this, 65% of syphilis tests are assigned an exposure category reported within 90 days, 80% within a year, and 87% within 2 years. These exposure categories are assigned through a hierarchy related to HIV risk in the following order: male-to-male sexual contact; heterosexual contact; “unknown HIV exposure” (no HIV risk factor reported [missing data]).

Statistical tests of trends over time used logistic regression with year as the independent variable. Odds ratios (ORs) and 97.5% confidence intervals (CI) are reported. When comparing two proportions, the 97.5% CI for the difference between the proportions are reported. The 97.5% confidence level was selected to account for the large population surveillance population sizes. All analyses were completed in SAS Enterprise Guide Version 8.4.

### Patient and Public Involvement statement

The work completed in this manuscript are considered routine public health practice and not research. As such, it was not appropriate or possible to involve patients or the public in the design, or conduct, or reporting, or dissemination plans of our work.

## RESULTS

Between 2017 and 2022, a total of 1,516,726 syphilis tests among males and 1,484,332 syphilis tests among females were included in the analysis. The proportion of tests that were positive for syphilis increased from 3.2% in 2017 to 6.6% in 2022 among males (OR 1.16, 97.5% CI 1.14-1.18) and from 0.5% in 2017 to 1.3% in 2022 among females (OR 1.21, 97.5% CI 1.15-1.27). Among females, the proportion of positive tests that were categorized as “current” infections increased from 19.0% in 2017 to 35.9% in 2022 (OR 1.22, 97.5% CI 1.17-1.28), whereas this decreased among males from 28.9% to 23.3% (OR 0.96, 97.5% CI 0.93-1.00). In 2020, the number of syphilis tests decreased by 27.9% among males and 24.6% among females, before recovering and surpassing 2019 counts in 2022.

### Non-positive syphilis tests

Between 2017 and 2022, the proportion of non-positive syphilis tests that had an HIV test on the same day +/-28 days increased from 87.7% to 92.7% among males (OR 1.14, 97.5% CI 1.08-1.19) and from 90.1% to 93.3% among females (OR 1.08, 97.5% CI 1.04-1.13, **Table 1**). In 2022, among males and females respectively, 90.7% and 91.5% had an HIV test on the same day, 92.7% and 93.3% had an HIV test on the same day +/-28 days, 93.4% and 93.8% on the same day +/-90 days, and 93.8% and 94.2% on the same day +/-180 days (**Table 2**). Between 2017 and 2022, 4.2% and 3.8% of non-positive tests had no corresponding HIV test regardless of time window among males and females, respectively (3.8% and 3.5% in 2022 alone).

**Table 1:** Number of syphilis tests, and number/proportion that had an HIV test on the same day +/-28 days, males and females, Ontario, Public Health Ontario, 2017-2022.

|  | Males |  | Females |  |
| --- | --- | --- | --- | --- |
|  | Total syphilis tests, N | Number/proportion that had an HIV test on the same day +/- 28 days, n (% [n/N]) | Total syphilis tests, N | Number/proportion that had an HIV test on the same day +/- 28 days, n (% [n/N]) |
| **Non-positive syphilis tests** |  |  |  |  |
| 2017 | 223,749 | 196,244 (87.7%) | 229,808 | 207,138 (90.1%) |
| 2018 | 250,420 | 224,380 (89.6%) | 256,726 | 236,174 (92.0%) |
| 2019 | 267,712 | 243,082 (90.8%) | 270,025 | 250,143 (92.6%) |
| 2020 | 191,064 | 176,202 (92.2%) | 203,238 | 188,933 (93.0%) |
| 2021 | 249,529 | 232,527 (93.2%) | 249,674 | 233,178 (93.4%) |
| 2022 | 262,800 | 243,590 (92.7%) | 262,899 | 245,327 (93.3%) |
| 2017-2022 | 1,445,274 | 1,316,025 (91.1%) | 1,472,370 | 1,360,893 (92.4%) |
| **All positive syphilis tests** |  |  |  |  |
| 2017 | 7,313 | 4,657 (63.7%) | 1,243 | 764 (61.5%) |
| 2018 | 9,236 | 6,327 (68.5%) | 1,517 | 948 (62.5%) |
| 2019 | 11,909 | 8,780 (73.7%) | 1,726 | 1,126 (65.2%) |
| 2020 | 10,662 | 8,184 (76.8%) | 1,599 | 1,032 (64.5%) |
| 2021 | 13,904 | 10,949 (78.7%) | 2,383 | 1,574 (66.1%) |
| 2022 | 18,428 | 14,492 (78.6%) | 3,494 | 2,351 (67.3%) |
| 2017-2022 | 71,452 | 53,389 (74.7%) | 11,962 | 7,795 (65.2%) |
| **“Current” positive syphilis tests** |  |  |  |  |
| 2017 | 2,113 | 1,212 (57.4%) | 236 | 131 (55.5%) |
| 2018 | 2,282 | 1,470 (64.4%) | 295 | 169 (57.3%) |
| 2019 | 2,905 | 1,997 (68.7%) | 406 | 249 (61.3%) |
| 2020 | 2,675 | 1,919 (71.7%) | 449 | 293 (65.3%) |
| 2021 | 3,388 | 2,505 (73.9%) | 831 | 603 (72.6%) |
| 2022 | 4,298 | 3,099 (72.1%) | 1,256 | 886 (70.5%) |
| 2017-2022 | 17,661 | 12,202 (69.1%) | 3,473 | 2,331 (67.1%) |
| **“Historical” positive syphilis tests** |  |  |  |  |
| 2017 | 5,200 | 3,445 (66.3%) | 1,007 | 633 (62.9%) |
| 2018 | 6,954 | 4,857 (69.8%) | 1,222 | 779 (63.7%) |
| 2019 | 9,004 | 6,783 (75.3%) | 1,320 | 877 (66.4%) |
| 2020 | 7,987 | 6,265 (78.4%) | 1,150 | 739 (64.3%) |
| 2021 | 10,516 | 8,444 (80.3%) | 1,552 | 971 (62.6%) |
| 2022 | 14,130 | 11,393 (80.6%) | 2,238 | 1,465 (65.5%) |
| 2017-2022 | 53,791 | 41,187 (76.6%) | 8,489 | 5,464 (64.4%) |

“Current” positive syphilis tests: Direct fluorescent antibody (DFA)-positive or a positive chemiluminescent microparticle immunoassay (CMIA) serology screen with a rapid plasma regain (RPR) antibody titre of $\geq 1:8$ were considered more likely to represent current (untreated) syphilis infections.; “Historical” positive syphilis tests: positive CMIA screen with an RPR $< 1:8$ were considered more likely to represent historical (treated/resolved) infections

**Table 2:** Number of syphilis tests, and number/proportion that had an HIV test on the same day, on the same day +/- 28 days, on the same day +/- 90 days, and on the same day +/- 180 days, by syphilis test result and year, males and females, Ontario, Public Health Ontario, 2022.

|  | Total<br>syphilis<br>tests<br>N | Number/proportion that<br>had an HIV test on the<br>same day<br>n (%) [n/N] | Number/proportion that<br>had an HIV test on the<br>same day +/- 28 days<br>n (%) [n/N] | Number/proportion that<br>had an HIV test on the<br>same day +/- 90 days<br>n (%) [n/N] | Number/proportion that<br>had an HIV test on the<br>same day +/- 180 days<br>n (%) [n/N] |
| --- | --- | --- | --- | --- | --- |
| **<u>Males</u>** |  |  |  |  |  |
| Non-positive syphilis tests | 262,800 | 238,376 (90.7%) | 243,590 (92.7%) | 245,333 (93.4%) | 246,502 (93.8%) |
| All positive syphilis tests | 18,428 | 13,078 (71.0%) | 14,492 (78.6%) | 15,571 (84.5%) | 16,255 (88.2%) |
| “Current” positive syphilis tests | 4,298 | 2,635 (61.3%) | 3,099 (72.1%) | 3,433 (79.9%) | 3,651 (84.9%) |
| “Historical” positive syphilis tests | 14,130 | 10,443 (73.9%) | 11,393 (80.6%) | 12,138 (85.9%) | 12,604 (89.2%) |
| **<u>Females</u>** |  |  |  |  |  |
| Non-positive syphilis tests | 262,899 | 240,500 (91.5%) | 245,327 (93.3%) | 246,669 (93.8%) | 247,576 (94.2%) |
| All positive syphilis tests | 3,494 | 2,029 (58.1%) | 2,351 (67.3%) | 2,641 (75.6%) | 2,865 (82.0%) |
| “Current” positive syphilis tests | 1,256 | 743 (59.2%) | 886 (70.5%) | 999 (79.5%) | 1,075 (85.6%) |
| “Historical” positive syphilis tests | 2,238 | 1,286 (57.5%) | 1,465 (65.5%) | 1,642 (73.4%) | 1,790 (80.0%) |

“Current” positive syphilis tests: Direct fluorescent antibody (DFA)-positive or a positive chemiluminescent microparticle immunoassay (CMIA) serology screen with a rapid plasma regain (RPR) antibody titre of $\geq 1:8$ were considered more likely to represent current (untreated) syphilis infections.; “Historical” positive syphilis tests: positive CMIA screen with an RPR $< 1:8$ were considered more likely to represent historical (treated/resolved) infections

### Positive syphilis tests

Between 2017 and 2022, among males, the proportion of positive syphilis tests that had an HIV test on the same day +/-28 days increased from 57.4% to 72.1% among “current” positives (OR 1.13, 97.5% CI 1.05-1.22), and from 66.3% to 80.6% among “historical” positives (OR 1.17, 97.5% CI 1.12-1.22, **Table 1**). “Current” positives were consistently less likely than “historical” positives to have had an HIV test on the same day +/-28 days (69.1% versus 76.6%, 97.5% CI [-0.084, -0.066]).

Among females, the proportion of “current” positives that had an HIV test on the same day +/-28 days increased from 55.5% in 2017 to 70.5% in 2022 (OR 1.16, 97.5% CI 1.11-1.22), but there was no significant time trend among “historical” positives (64.4% overall, **Table 1**). Unlike in males, “current” positives were more likely than “historical” positives to have had an HIV test on the same day +/-28 days overall (67.1% versus 64.4%, 97.5% CI [0.0062, 0.049]), but this varied by year. Between 2017-2022 cumulatively, a larger proportion of syphilis tests among males had an HIV test on the same day +/-28 days compared to females for both “current” (69.1% versus 67.1%, 97.5% CI [0.00023, 0.039]) and “historical” positive syphilis tests (76.6% versus 64.4%, 97.5% CI [0.11, 0.13]) (**Table 1**). Between 2017 and 2022, 5.5% and 7.3% of all positive syphilis tests among males and females, respectively, had no corresponding HIV test regardless of time window (4.9% and 6.0% in 2022 alone). For a detailed breakdown of the results, refer to Supplemental Tables 1-2.

### HIV diagnoses with positive syphilis tests

Among females between 2017 and 2022, 0.3% of all positive syphilis tests with an HIV test on the same day +/-28 days had a positive HIV test result, and there was no difference between “current” and “historical” positive syphilis tests (97.5% CI [-0.0032, 0.0029], **Table 3**). Among males however, 1.2% of all positive syphilis tests with an HIV test on the same day +/-28 days had a positive HIV test result, and this differed greatly by category of positive syphilis test result – 2.7% among “current” positives and 0.8% among “historical” positives (97.5% CI [0.016, 0.023], **Table 3**). These proportions both decreased over time (OR 0.82, 97.5% CI 0.77-0.88 for “current”; OR 0.85, 97.5% CI 0.79-0.92 for “historical”, **Supplemental Table 4**), as the number of positive syphilis tests (denominator) increased for both. When examining within HIV exposure categories, 1.5% of “current” and 0.5% and “historical” positive syphilis tests with an HIV test on the same day +/-28 days had a positive HIV test result among males reporting heterosexual contact (97.5% CI [0.004, 0.017], **Table 3**), compared to 2.9% and 0.7% respectively among males reporting male-to-male sexual contact (97.5% CI [0.018, 0.027]), and 3.4% and 1.5% among males with unknown HIV exposure (97.5% CI [0.0092, 0.029]). For a detailed breakdown of the results, refer to Supplemental Tables 3-4.

**Table 3:** Number of positive syphilis tests that had an HIV test on the same day +/- 28 days, and number/proportion that had a positive HIV test result, by sex, HIV exposure category (among males), and syphilis test result, Ontario, Public Health Ontario, 2017-2022 (cumulative)

|  | Positive syphilis tests<br>with an HIV test* | Number/proportion that<br>had a positive HIV test<br>result |
| --- | --- | --- |
|  | N | n (% [n/N]) |
| **All positive syphilis tests** |  |  |
| **Females** | 7,795 | 24 (0.3%) |
| **Males (all)** | 53,389 | 665 (1.2%) |
| **Males with male-to-male sexual contact** | 38,589 | 450 (1.2%) |
| **Males with heterosexual contact** | 6,226 | 50 (0.8%) |
| **Males with unknown HIV exposure** | 7,918 | 159 (2.0%) |
| **“Current” positive syphilis tests** |  |  |
| **Females** | 2,331 | 7 (0.3%) |
| **Males (all)** | 12,202 | 334 (2.7%) |
| **Males with male-to-male sexual contact** | 7,863 | 231 (2.9%) |
| **Males with heterosexual contact** | 2,070 | 31 (1.5%) |
| **Males with unknown HIV exposure** | 1,981 | 68 (3.4%) |
| **“Historical” positive syphilis tests** |  |  |
| **Females** | 5,464 | 17 (0.3%) |
| **Males (all)** | 41,187 | 331 (0.8%) |
| **Males with male-to-male sexual contact** | 30,726 | 219 (0.7%) |
| **Males with heterosexual contact** | 4,156 | 19 (0.5%) |
| **Males with unknown HIV exposure** | 5,937 | 91 (1.5%) |

“Current” positive syphilis tests: Direct fluorescent antibody (DFA)-positive or a positive chemiluminescent microparticle immunoassay (CMIA) serology screen with a rapid plasma regain (RPR) antibody titre of $\geq 1:8$ were considered more likely to represent current (untreated) syphilis infections.; “Historical” positive syphilis tests: positive CMIA screen with an RPR $< 1:8$ were considered more likely to represent historical (treated/resolved) infections. \*HIV test on the same day +/- 28 days.

## DISCUSSION

Our results provide insight into observance of HIV testing guidelines for individuals being tested for syphilis in Ontario. It is encouraging that approximately 9 in 10 non-positive syphilis tests had a HIV test on the same day for both males and females, and since non-positive tests make up the vast majority (>93%) of all syphilis tests, it can be concluded that most syphilis tests in general are indeed accompanied by HIV tests among individuals with no evidence of a previous HIV diagnosis. In contrast, only ∼3/4 of all positive syphilis tests among males and ∼2/3 among females had an HIV test on the same day +/-28 days. Furthermore, only ∼7/10 “current” positive syphilis tests among males had an HIV test on the same day +/-28 days. One explanation of individuals with positive syphilis tests being less likely to receive timely HIV testing may pertain to these tests representing diagnostic testing among individuals with symptoms and/or known exposures, where providers may be particularly focused on syphilis diagnosis.

Further, some of these positive syphilis tests will represent repeat or follow-up tests after diagnosis and treatment of syphilis to monitor treatment progress, where concurrent HIV testing may not have been indicated, or conversely may have in fact been opportune. It is encouraging that larger proportions of positive syphilis tests did have corresponding HIV tests when applying a larger time window (+/-180 days). Non-positive syphilis tests may be conducted as a part of comprehensive STI screening among asymptomatic individuals, including those on HIV pre-exposure prophylaxis (PrEP). Providers have previously reported that they may not routinely offer HIV testing due to policy barriers, feeling rushed with patients, fear of offending their patients, or the perception that their patient is low risk (18) (19).

Where HIV testing was performed, individuals positive for syphilis were more likely to be diagnosed as HIV-positive compared to the general population testing for HIV, highlighting the pertinence of this population in identifying undiagnosed individuals living with HIV. In 2022, 0.4% of females positive for syphilis in our study were diagnosed with HIV, compared to the general population HIV test positivity of 0.05% among females (p<0.05) (20). This significant difference (p<0.05 for all) was consistent for males with male-to-male sexual contact (0.8% versus 0.31% respectively); heterosexual contact (0.8% versus 0.05%); and unknown HIV exposure (1.6% versus 0.13%). These differences were greater among “current” positive syphilis tests. These results provide further support to the established correlation between syphilis and HIV infections (4), especially among GBMSM (9) (10) (11).

Many of the risk factors for STIs, including syphilis, overlap with risk factors for HIV and would therefore indicate testing for both (21). Accurate assessment of a patient’s risk for HIV can be difficult because patients may hesitate to disclose for fear of stigmatization or may self-perceive their behaviours to be low risk for HIV acquisition (22). For this reason, the United States CDC guidelines state that HIV screening should accompany STI testing, “regardless of whether the patient reports any specific behavioral risks for HIV” (8). However, a large dataset from emergency department visits for STI testing in the United States between 2010 and 2018 found that only 1.5% of these encounters included ordering of both syphilis and HIV testing (15). Our findings showed that among positive syphilis tests with HIV tests on the same day +/-28 days, males that reported heterosexual contact (and therefore no male-to-male sexual contact by virtue of the exposure category hierarchy) were more likely to receive positive HIV test results than their female counterparts (p<0.05). This was an interesting finding in that one might expect more comparable HIV risk with shared heterosexual exposure. Also, males with unknown HIV exposure, who may not have disclosed or been asked about their HIV risk factors, were the most likely to receive positive HIV test results. This dearth in disclosure can preclude sexual health counselling and provision of prevention options, including PrEP. Our results also demonstrate potential limitations of such self-reported data in clinical settings and provoke questions of how offering testing based only on self-reported risk of the patient may contribute to missed diagnoses and ongoing transmission. This is tempered however with consideration that risk factors may indeed be discussed but not consistently documented on test requisition forms.

Our analysis is not without limitations. First, individuals’ HIV tests may not always link to their syphilis tests. This has implications for [1] identifying people previously diagnosed with HIV (and exclusion of their syphilis tests); and [2] identifying corresponding HIV tests. Although we employed linkage based on health card numbers (a unique ID for most Ontarians), dates of birth, and first and last names, failed linkages could result from incomplete or inconsistent data, or non-nominal HIV testing. The majority of anonymous HIV testing in Ontario is conducted in males (20), therefore males may be more likely to have no linked HIV test. Anonymous testing comprised a relatively small proportion (between 0.8 and 2.4% per year) of HIV tests between 2017 and 2022. Second, given that we were limited to syphilis laboratory testing data alone, we could not distinguish current syphilis infections from treated/resolved infections. Our “current” definition (i.e. RPR ≥1:8) is a conservative estimate of infectious syphilis that was based on national guidelines and consultation with health care providers with syphilis expertise (5). Classification of syphilis test results based on longitudinal analyses was not employed due to the need for historical data (dataset restricted to 6 years), limitations pertaining to patient linkage to enable validation of the longitudinal analysis using case and clinical information, and the preference for more simplistic methods intended for surveillance. Our categorization based on RPR titres is not a validated method and will encompass misclassification on the individual level; however, we believe it to be reasonable for population-level surveillance purposes. To support this, the proportion of individuals with an RPR ≥1:8 among individuals tested was 0.97%, which was comparable to the proportion of infectious syphilis cases (identified through public health reporting) among individuals tested (0.71%). The main objective of this analysis was to find opportunities for public health intervention to improve testing for HIV and we reasoned that any syphilis testing visit could be an opportunity for HIV testing. The rate of syphilis testing in our dataset is approximately 1.16 tests per person per year, which aligns with the recommendation that individuals at risk for HIV be screened at least annually Lastly, we had incomplete data with respect to HIV exposure category, and these data are subject to recall and social desirability biases, as discussed. This may affect interpretations among exposure categories, especially as males with unknown HIV exposure had the largest proportions with positive HIV test results.

Since most syphilis and HIV testing are centralized at PHO and our analyses used all tests in the study period, our results are representative provincially; however, they may not be generalized to other jurisdictions.

## CONCLUSIONS

In conclusion, most individuals who were tested for syphilis in Ontario between 2017 and 2022 were also tested for HIV. Individuals with positive syphilis tests, however, despite some important caveats, were less likely to be tested for HIV compared to individuals non-positive for syphilis, representing a potential opportunity for enhanced HIV testing among those positive for syphilis in accordance with HIV testing guidelines. When tested for HIV, individuals positive for syphilis were considerably more likely to be diagnosed as HIV-positive compared to the general population testing for HIV, especially males and in particular males with “current” positive syphilis test results. Ensuring that individuals with positive syphilis results are tested for HIV in a timely manner may identify people living with previously undiagnosed HIV.

## Data Availability Statement

All available data are presented in this manuscript.

## Ethics Statements

This project did not require research ethics committee approval as the activities described in this manuscript are considered routine public health practice and not research. These activities were conducted in fulfillment of Public Health Ontario’s legislated mandate “to provide scientific and technical advice and support to the health care system and the Government of Ontario in order to protect and promote the health of Ontarians…” (Ontario Agency for Health Protection and Promotion Act, SO 2007, c 10, Schedule K).

## Funding Statement

This work was completed as part of routine operations for Public Health Ontario. This research received no specific grant from any funding agency in the public, commercial or not-for-profit sectors.

## Competing Interests

There are no competing interests for authors.

## Acknowledgments

The authors would like to acknowledge Ken English, HIV & Hepatitis C Programs at the Ontario Ministry of Health for the data request that lead to this analysis. We also acknowledge all staff of PHO’s laboratory. PHO’s laboratory and offices are situated on the traditional territory of many nations including the Mississaugas of the Credit, the Anishnabeg, the Chippewa, the Haudenosaunee and the Wendat peoples and is now home to many diverse First Nations, Inuit and Métis peoples.

## Author’s Contribution

Each author meets all four of the ICMJE criteria outlined for authorship.

## Notes

### Competing Interest Statement

The authors have declared no competing interest.

### Author Declarations

Ethics committee/IRB of Public Health Ontario waived ethical approval for this work.

