## Supplemental Material for "How often did syphilis tests have corresponding HIV tests in Ontario, Canada? A retrospective analysis of comprehensive laboratory data"

**Supplementary Materials**

**Table 1: Number of syphilis tests, and number/proportion that had an HIV test on the same day, on the same day +/- 28 days, on the same day +/- 90 days, and on the same day +/- 180 days, by syphilis test result and year, males, Ontario, Public Health Ontario, 2017-2022**

|  | Total syphilis tests | Number/proportion that had an HIV test on the same day | Number/proportion that had an HIV test on the same day +/- 28 days | Number/proportion that had an HIV test on the same day +/- 90 days | Number/proportion that had an HIV test on the same day +/- 180 days |
| --- | --- | --- | --- | --- | --- |
|  | **N** | **n (% [n/N])** | **n (% [n/N])** | **n (% [n/N])** | **n (% [n/N])** |
| Non-positive syphilis tests | |  |  |  |  |
| 2017 | 223,749 | 191,792 (85.7%) | 196,244 (87.7%) | 198,364 (88.7%) | 200,321 (89.5%) |
| 2018 | 250,420 | 219,409 (87.6%) | 224,380 (89.6%) | 226,556 (90.5%) | 228,563 (91.3%) |
| 2019 | 267,712 | 238,441 (89.1%) | 243,082 (90.8%) | 245,326 (91.6%) | 247,196 (92.3%) |
| 2020 | 191,064 | 172,818 (90.5%) | 176,202 (92.2%) | 177,497 (92.9%) | 178,569 (93.5%) |
| 2021 | 249,529 | 230,086 (92.2%) | 232,527 (93.2%) | 234,154 (93.8%) | 235,321 (94.3%) |
| 2022 | 262,800 | 238,376 (90.7%) | 243,590 (92.7%) | 245,333 (93.4%) | 246,502 (93.8%) |
| 2017-2022 | 1,445,274 | 1,290,922 (89.3%) | 1,316,025 (91.1%) | 1,327,230 (91.8%) | 1,336,472 (92.5%) |
| All positive syphilis tests | |  |  |  |  |
| 2017 | 7,313 | 3,992 (54.6%) | 4,657 (63.7%) | 5,234 (71.6%) | 5,631 (77.0%) |
| 2018 | 9,236 | 5,606 (60.7%) | 6,327 (68.5%) | 6,995 (75.7%) | 7,445 (80.6%) |
| 2019 | 11,909 | 7,882 (66.2%) | 8,780 (73.7%) | 9,585 (80.5%) | 10,101 (84.8%) |
| 2020 | 10,662 | 7,350 (68.9%) | 8,184 (76.8%) | 8,830 (82.8%) | 9,300 (87.2%) |
| 2021 | 13,904 | 10,100 (72.6%) | 10,949 (78.7%) | 11,798 (84.9%) | 12,320 (88.6%) |
| 2022 | 18,428 | 13,078 (71.0%) | 14,492 (78.6%) | 15,571 (84.5%) | 16,255 (88.2%) |
| 2017-2022 | 71,452 | 48,008 (67.2%) | 53,389 (74.7%) | 58,013 (81.2%) | 61,052 (85.4%) |
| “Current” positive syphilis tests | | |  |  |  |
| 2017 | 2,113 | 963 (45.6%) | 1,212 (57.4%) | 1,437 (68.0%) | 1,585 (75.0%) |
| 2018 | 2,282 | 1,204 (52.8%) | 1,470 (64.4%) | 1,649 (72.3%) | 1,801 (78.9%) |
| 2019 | 2,905 | 1,641 (56.5%) | 1,997 (68.7%) | 2,257 (77.7%) | 2,401 (82.7%) |
| 2020 | 2,675 | 1,557 (58.2%) | 1,919 (71.7%) | 2,142 (80.1%) | 2,298 (85.9%) |
| 2021 | 3,388 | 2,154 (63.6%) | 2,505 (73.9%) | 2,778 (82.0%) | 2,933 (86.6%) |
| 2022 | 4,298 | 2,635 (61.3%) | 3,099 (72.1%) | 3,433 (79.9%) | 3,651 (84.9%) |
| 2017-2022 | 17,661 | 10,154 (57.5%) | 12,202 (69.1%) | 13,696 (77.5%) | 14,669 (83.1%) |
| “Historical” positive syphilis tests | | |  |  |  |
| 2017 | 5,200 | 3,029 (58.3%) | 3,445 (66.3%) | 3,797 (73.0%) | 4,046 (77.8%) |
| 2018 | 6,954 | 4,402 (63.3%) | 4,857 (69.8%) | 5,346 (76.9%) | 5,644 (81.2%) |
| 2019 | 9,004 | 6,241 (69.3%) | 6,783 (75.3%) | 7,328 (81.4%) | 7,700 (85.5%) |
| 2020 | 7,987 | 5,793 (72.5%) | 6,265 (78.4%) | 6,688 (83.7%) | 7,002 (87.7%) |
| 2021 | 10,516 | 7,946 (75.6%) | 8,444 (80.3%) | 9,020 (85.8%) | 9,387 (89.3%) |
| 2022 | 14,130 | 10,443 (73.9%) | 11,393 (80.6%) | 12,138 (85.9%) | 12,604 (89.2%) |
| 2017-2022 | 53,791 | 37,854 (70.4%) | 41,187 (76.6%) | 44,317 (82.4%) | 46,383 (86.2%) |

“Current” positive syphilis tests: Direct fluorescent antibody (DFA)-positive or a positive chemiluminescent microparticle immunoassay (CMIA) serology screen with a rapid plasma regain (RPR) antibody titre of ≥ 1:8 were considered more likely to represent current (untreated) syphilis infections.; “Historical” positive syphilis tests: positive CMIA screen with an RPR < 1:8 were considered more likely to represent historical (treated/resolved) infections.

**Table 2: Number of syphilis tests, and number/proportion that had an HIV test on the same day, on the same day +/- 28 days, on the same day +/- 90 days, and on the same day +/- 180 days, by syphilis test result and year, females, Ontario, Public Health Ontario, 2017-2022**

|  | Total syphilis tests | Number/proportion that had an HIV test on the same day | Number/proportion that had an HIV test on the same day +/- 28 days | Number/proportion that had an HIV test on the same day +/- 90 days | Number/proportion that had an HIV test on the same day +/- 180 days |
| --- | --- | --- | --- | --- | --- |
|  | **N** | **n (% [n/N])** | **n (% [n/N])** | **n (% [n/N])** | **n (% [n/N])** |
| Non-positive syphilis tests | |  |  |  |  |
| 2017 | 229,808 | 203,325 (88.5%) | 207,138 (90.1%) | 208,478 (90.7%) | 209,845 (91.3%) |
| 2018 | 256,726 | 231,843 (90.3%) | 236,174 (92.0%) | 237,606 (92.6%) | 238,925 (93.1%) |
| 2019 | 270,025 | 246,328 (91.2%) | 250,143 (92.6%) | 251,507 (93.1%) | 252,706 (93.6%) |
| 2020 | 203,238 | 185,794 (91.4%) | 188,933 (93.0%) | 189,933 (93.5%) | 190,757 (93.9%) |
| 2021 | 249,674 | 231,209 (92.6%) | 233,178 (93.4%) | 234,404 (93.9%) | 235,366 (94.3%) |
| 2022 | 262,899 | 240,500 (91.5%) | 245,327 (93.3%) | 246,669 (93.8%) | 247,576 (94.2%) |
| 2017-2022 | 1,472,370 | 1,338,999 (90.9%) | 1,360,893 (92.4%) | 1,368,597 (93.0%) | 1,375,175 (93.4%) |
| All positive syphilis tests | |  |  |  |  |
| 2017 | 1,243 | 635 (51.1%) | 764 (61.5%) | 841 (67.7%) | 901 (72.5%) |
| 2018 | 1,517 | 837 (55.2%) | 948 (62.5%) | 1,097 (72.3%) | 1,174 (77.4%) |
| 2019 | 1,726 | 982 (56.9%) | 1,126 (65.2%) | 1,275 (73.9%) | 1,367 (79.2%) |
| 2020 | 1,599 | 880 (55.0%) | 1,032 (64.5%) | 1,172 (73.3%) | 1,264 (79.0%) |
| 2021 | 2,383 | 1,372 (57.6%) | 1,574 (66.1%) | 1,814 (76.1%) | 1,967 (82.5%) |
| 2022 | 3,494 | 2,029 (58.1%) | 2,351 (67.3%) | 2,641 (75.6%) | 2,865 (82.0%) |
| 2017-2022 | 11,962 | 6,735 (56.3%) | 7,795 (65.2%) | 8,840 (73.9%) | 9,538 (79.7%) |
| “Current” positive syphilis tests | | |  |  |  |
| 2017 | 236 | 104 (44.1%) | 131 (55.5%) | 152 (64.4%) | 172 (72.9%) |
| 2018 | 295 | 142 (48.1%) | 169 (57.3%) | 200 (67.8%) | 225 (76.3%) |
| 2019 | 406 | 209 (51.5%) | 249 (61.3%) | 289 (71.2%) | 321 (79.1%) |
| 2020 | 449 | 240 (53.5%) | 293 (65.3%) | 340 (75.7%) | 369 (82.2%) |
| 2021 | 831 | 499 (60.0%) | 603 (72.6%) | 680 (81.8%) | 730 (87.8%) |
| 2022 | 1,256 | 743 (59.2%) | 886 (70.5%) | 999 (79.5%) | 1,075 (85.6%) |
| 2017-2022 | 3,473 | 1,937 (55.8%) | 2,331 (67.1%) | 2,660 (76.6%) | 2,892 (83.3%) |
| “Historical” positive syphilis tests | | |  |  |  |
| 2017 | 1,007 | 531 (52.7%) | 633 (62.9%) | 689 (68.4%) | 729 (72.4%) |
| 2018 | 1,222 | 695 (56.9%) | 779 (63.7%) | 897 (73.4%) | 949 (77.7%) |
| 2019 | 1,320 | 773 (58.6%) | 877 (66.4%) | 986 (74.7%) | 1,046 (79.2%) |
| 2020 | 1,150 | 640 (55.7%) | 739 (64.3%) | 832 (72.3%) | 895 (77.8%) |
| 2021 | 1,552 | 873 (56.3%) | 971 (62.6%) | 1,134 (73.1%) | 1,237 (79.7%) |
| 2022 | 2,238 | 1,286 (57.5%) | 1,465 (65.5%) | 1,642 (73.4%) | 1,790 (80.0%) |
| 2017-2022 | 8,489 | 4,798 (56.5%) | 5,464 (64.4%) | 6,180 (72.8%) | 6,646 (78.3%) |

“Current” positive syphilis tests: Direct fluorescent antibody (DFA)-positive or a positive chemiluminescent microparticle immunoassay (CMIA) serology screen with a rapid plasma regain (RPR) antibody titre of ≥ 1:8 were considered more likely to represent current (untreated) syphilis infections.; “Historical” positive syphilis tests: positive CMIA screen with an RPR < 1:8 were considered more likely to represent historical (treated/resolved) infections.

**Table 3: Number of positive syphilis tests that had an HIV test on the same day +/- 28 days, and number/proportion that had a positive HIV test result, by sex, syphilis test result and year, Ontario, Public Health Ontario, 2017-2022**

|  | **Overall** | | **Females** | | **Males** | |
| --- | --- | --- | --- | --- | --- | --- |
|  | **Positive syphilis tests with an HIV test*** | **Number/proportion that had a positive HIV test result** | **Positive syphilis tests with an HIV test*** | **Number/proportion that had a positive HIV test result** | **Positive syphilis tests with an HIV test*** | **Number/proportion that had a positive HIV test result** |
|  | **N** | **n (% [n/N])** | **N** | **n (% [n/N])** | **N** | **n (% [n/N])** |
| **All positive syphilis tests** | |  |  |  |  |  |
| **2017** | 5,463 | 118 (2.2%) | 764 | 3 (0.4%) | 4,657 | 115 (2.5%) |
| **2018** | 7,331 | 103 (1.4%) | 948 | 2 (0.2%) | 6,327 | 101 (1.6%) |
| **2019** | 9,976 | 122 (1.2%) | 1,126 | 5 (0.4%) | 8,780 | 116 (1.3%) |
| **2020** | 9,297 | 91 (1.0%) | 1,032 | 2 (0.2%) | 8,184 | 89 (1.1%) |
| **2021** | 12,638 | 116 (0.9%) | 1,574 | 3 (0.2%) | 10,949 | 111 (1.0%) |
| **2022** | 17,024 | 144 (0.8%) | 2,351 | 9 (0.4%) | 14,492 | 133 (0.9%) |
| **2017-2022** | 61,729 | 694 (1.1%) | 7,795 | 24 (0.3%) | 53,389 | 665 (1.2%) |
| **“Current” positive syphilis tests** | | |  |  |  |  |
| **2017** | 1,350 | 65 (4.8%) | 131 | 0 (0.0%) | 1,212 | 65 (5.4%) |
| **2018** | 1,653 | 49 (3.0%) | 169 | 0 (0.0%) | 1,470 | 49 (3.3%) |
| **2019** | 2,268 | 57 (2.5%) | 249 | 1 (0.4%) | 1,997 | 55 (2.8%) |
| **2020** | 2,228 | 52 (2.3%) | 293 | 1 (0.3%) | 1,919 | 51 (2.7%) |
| **2021** | 3,142 | 58 (1.8%) | 603 | 0 (0.0%) | 2,505 | 57 (2.3%) |
| **2022** | 4,031 | 63 (1.6%) | 886 | 5 (0.6%) | 3,099 | 57 (1.8%) |
| **2017-2022** | 14,672 | 344 (2.3%) | 2,331 | 7 (0.3%) | 12,202 | 334 (2.7%) |
| **“Historical” positive syphilis tests** | | |  |  |  |  |
| **2017** | 4,113 | 53 (1.3%) | 633 | 3 (0.5%) | 3,445 | 50 (1.5%) |
| **2018** | 5,678 | 54 (1.0%) | 779 | 2 (0.3%) | 4,857 | 52 (1.1%) |
| **2019** | 7,708 | 65 (0.8%) | 877 | 4 (0.5%) | 6,783 | 61 (0.9%) |
| **2020** | 7,069 | 39 (0.6%) | 739 | 1 (0.1%) | 6,265 | 38 (0.6%) |
| **2021** | 9,496 | 58 (0.6%) | 971 | 3 (0.3%) | 8,444 | 54 (0.6%) |
| **2022** | 12,993 | 81 (0.6%) | 1,465 | 4 (0.3%) | 11,393 | 76 (0.7%) |
| **2017-2022** | 47,057 | 350 (0.7%) | 5,464 | 17 (0.3%) | 41,187 | 331 (0.8%) |

“Current” positive syphilis tests: Direct fluorescent antibody (DFA)-positive or a positive chemiluminescent microparticle immunoassay (CMIA) serology screen with a rapid plasma regain (RPR) antibody titre of ≥ 1:8 were considered more likely to represent current (untreated) syphilis infections.; “Historical” positive syphilis tests: positive CMIA screen with an RPR < 1:8 were considered more likely to represent historical (treated/resolved) infections. *HIV test on the same day +/- 28 days.

**Table 4: Number of positive syphilis tests that had an HIV test on the same day +/- 28 days, and number/proportion that had a positive HIV test result, by HIV exposure category, syphilis test result and year, males, Ontario, Public Health Ontario, 2017-2022**

|  | **Males with male-to-male sexual contact** | | **Males with heterosexual contact** | | **Males with unknown HIV exposure** | |
| --- | --- | --- | --- | --- | --- | --- |
|  | **Positive syphilis tests with an HIV test*** | **Number/proportion that had a positive HIV test result** | **Positive syphilis tests with an HIV test*** | **Number/proportion that had a positive HIV test result** | **Positive syphilis tests with an HIV test*** | **Number/proportion that had a positive HIV test result** |
|  | **N** | **n (% [n/N])** | **N** | **n (% [n/N])** | **N** | **n (% [n/N])** |
| **All positive syphilis tests** | |  |  |  |  |  |
| **2017** | 3,193 | 82 (2.6%) | 683 | 12 (1.8%) | 753 | 21 (2.8%) |
| **2018** | 4,496 | 65 (1.4%) | 770 | 7 (0.9%) | 1,004 | 28 (2.8%) |
| **2019** | 6,508 | 77 (1.2%) | 995 | 5 (0.5%) | 1,193 | 32 (2.7%) |
| **2020** | 6,236 | 67 (1.1%) | 886 | 3 (0.3%) | 966 | 19 (2.0%) |
| **2021** | 8,038 | 80 (1.0%) | 1,241 | 10 (0.8%) | 1,511 | 19 (1.3%) |
| **2022** | 10,118 | 79 (0.8%) | 1,651 | 13 (0.8%) | 2,491 | 40 (1.6%) |
| **2017-2022** | 38,589 | 450 (1.2%) | 6,226 | 50 (0.8%) | 7,918 | 159 (2.0%) |
| **“Current” positive syphilis tests** | | |  |  |  |  |
| **2017** | 874 | 44 (5.0%) | 180 | 9 (5.0%) | 146 | 12 (8.2%) |
| **2018** | 1,071 | 35 (3.3%) | 217 | 2 (0.9%) | 161 | 11 (6.8%) |
| **2019** | 1,427 | 41 (2.9%) | 294 | 2 (0.7%) | 241 | 11 (4.6%) |
| **2020** | 1,319 | 42 (3.2%) | 302 | 3 (1.0%) | 262 | 6 (2.3%) |
| **2021** | 1,527 | 34 (2.2%) | 463 | 9 (1.9%) | 439 | 13 (3.0%) |
| **2022** | 1,645 | 35 (2.1%) | 614 | 6 (1.0%) | 732 | 15 (2.0%) |
| **2017-2022** | 7,863 | 231 (2.9%) | 2,070 | 31 (1.5%) | 1,981 | 68 (3.4%) |
| **“Historical” positive syphilis tests** | | |  |  |  |  |
| **2017** | 2,319 | 38 (1.6%) | 503 | 3 (0.6%) | 607 | 9 (1.5%) |
| **2018** | 3,425 | 30 (0.9%) | 553 | 5 (0.9%) | 843 | 17 (2.0%) |
| **2019** | 5,081 | 36 (0.7%) | 701 | 3 (0.4%) | 952 | 21 (2.2%) |
| **2020** | 4,917 | 25 (0.5%) | 584 | 0 (0.0%) | 704 | 13 (1.8%) |
| **2021** | 6,511 | 46 (0.7%) | 778 | 1 (0.1%) | 1,072 | 6 (0.6%) |
| **2022** | 8,473 | 44 (0.5%) | 1,037 | 7 (0.7%) | 1,759 | 25 (1.4%) |
| **2017-2022** | 30,726 | 219 (0.7%) | 4,156 | 19 (0.5%) | 5,937 | 91 (1.5%) |

“Current” positive syphilis tests: Direct fluorescent antibody (DFA)-positive or a positive chemiluminescent microparticle immunoassay (CMIA) serology screen with a rapid plasma regain (RPR) antibody titre of ≥ 1:8 were considered more likely to represent current (untreated) syphilis infections.; “Historical” positive syphilis tests: positive CMIA screen with an RPR < 1:8 were considered more likely to represent historical (treated/resolved) infections. *HIV test on the same day +/- 28 days.
